# Empirical Evidence for Cognitive Subgroups in Body Dysmorphic Disorder

**DOI:** 10.1101/2020.06.29.20143073

**Authors:** Amy Malcolm, Sarah N. Brennan, Sally A. Grace, Toni D. Pikoos, Wei Lin Toh, Izelle Labuschagne, Ben Buchanan, Ryan A. Kaplan, David J. Castle, Susan L. Rossell

**Affiliations:** Centre for Mental Health, Faculty of Health, Arts & Design, Swinburne University of Technology, Hawthorn, VIC, Australia; The Royal Children’s Hospital, Melbourne, VIC, Australia; School of Behavioural and Health Sciences, Australian Catholic University, Melbourne, VIC, Australia; Sydney Body Dysmorphic Disorder & Body Image Clinic, Bondi Junction, NSW, Australia; Department of Mental Health, St Vincent’s Hospital, Melbourne, VIC, Australia; The University of Melbourne, Melbourne, VIC, Australia

**Keywords:** BDD, body dysmorphic disorder, cognition, neuropsychology, neurocognitive

## Abstract

**Objective:** Current understanding of cognitive functioning in body dysmorphic disorder (BDD) is limited, owing to few studies, small sample sizes, and assessment across only limited cognitive domains. Existing research has also shown inconsistent findings, with both intact and impaired cognition reported in BDD, which might point toward cognitive heterogeneity in the disorder. This study aimed to examine the cognitive profile of BDD in a large sample across eight cognitive domains, and to explore whether cognitive subgroups might be identified within BDD.

**Methods:** Cognitive domains of inhibition/flexibility, working memory, speed of processing, reasoning and problem-solving, visual and verbal learning, attention/vigilance and social cognition were assessed and compared between 65 BDD patients and 70 healthy controls. Then, hierarchical clustering analysis was conducted on the BDD group’s cognitive data.

**Results:** Group-average comparisons demonstrated significantly poorer cognitive functioning in BDD than healthy controls in all domains except for attention/vigilance and social cognition. Cluster analysis identified two divergent cognitive subgroups within our BDD cohort characterised by *i*) broadly intact cognitive function with mild selective impairments (72.3%), and *ii)* broadly impaired cognitive function (27.7%). However, the clusters did not significantly differ on clinical parameters or most sociodemographic characteristics.

**Conclusion:** Cognitively diverse subgroups were identified within BDD, yet these were obscured in group-average comparisons. However, subgroup profiles of cognitive functioning seem unrelated to the clinical presentation of BDD. Further research into the underlying mechanisms of cognition in BDD is warranted.

**Significant Outcomes:**

- Cognitive function varies widely among people with body dysmorphic disorder (BDD) and includes subgroups characterised by broadly intact or broadly impaired cognitive profiles.
- However, reduced performances in visual learning and working memory as relative to healthy participants were identified across both BDD cognitive subgroups to differing degrees.
- Clinical characteristics were not significantly different among the two BDD cognitive subgroups; what role cognitive functioning may play in the aetiology or presentation of BDD remains unclear.

**Limitations:**

- The output of clustering analysis is sensitive to the choice of variables entered and requires replication with analogous measures of cognition.
- Psychiatric comorbidities and medications could have impacted cognitive performances in the BDD group in an unaccounted-for manner.

**Data availability statement:** Author elects to not share data.

---

Body dysmorphic disorder (BDD) is a complex psychiatric condition that affects approximately 2% of the population (1) and is associated with high rates of suicidal behaviour, hospitalisation and severe psychosocial impairment (2, 3). Despite the potentially devastating impact of BDD, the disorder remains relatively understudied with regards to underlying mechanisms of development. One area requiring further research is cognitive functioning in BDD, particularly as deficits in specific functions such as memory, visual processes, reasoning, and social cognition are theorised to be important in the development and/or maintenance of the disorder (4).

There has been limited investigation of the general cognitive profile of BDD. Using the Repeatable Battery for the Assessment of Neuropsychological Status (RBANS), Toh, Castle and Rossell (5) found significantly poorer overall performance in BDD (*n* = 21) than in healthy controls (*n* = 21), that appeared to be driven by significantly poorer performance in attention and immediate memory in BDD. No significant differences between BDD and healthy control participants were found in visuospatial construction, language, or delayed memory. However, the authors noted that ceiling performance effects might have obscured further group differences in these domains (5). Nevertheless, of the handful of other studies that have examined cognitive functioning in BDD, several have found significantly reduced performance relative to healthy controls in functions related to planning, flexibility, inhibition, visual organisation, working memory and processing speed (6-10). However, findings related to visual recall, verbal recall and fluency have been inconsistent in demonstrating either intact or significantly reduced performance in BDD relative to healthy controls (6-9, 11). These inconsistent findings might relate to study differences in test selection, sample characteristics, or reduced statistical power due to generally modest sample sizes in these studies (i.e., *n* per group *=* 14 to 21). Thus, an examination of broad cognitive functioning conducted in a large sample of BDD participants would assist in clarifying our understanding of cognition in the disorder.

Another possible explanation for the inconsistencies in published studies might be that there is no unitary cognitive profile associated with BDD. Rather, individuals with the disorder might be better characterised according to subgroups falling along a spectrum of cognitive functioning ranging from intact to markedly impaired. The presence of such subgroups would be obscured in comparisons based on group-averaged scores, with mean scores trending toward the most dominant subgroup profiles. Thus, inconsistent findings across cognitive studies in BDD could reflect the sampling of disparate cognitive subgroups. The presence of such cognitive subgroups has been established in other psychiatric disorders such as bipolar disorder and schizophrenia (12), and therefore may also exist in BDD. If such subgroups are present within BDD, their identification will facilitate a nuanced understanding of cognitive deficits in the disorder, which can be used to identify clinical or biological correlates that may be important for the development of appropriate treatments for BDD and for understanding its aetiological processes. For example, specific cognitive profiles might be associated with differences in illness prognosis or treatment outcomes, which in turn could inform the potential value of adjunctive cognitive remediation approaches for specific individuals.

## Aims of the study

The present study had two aims. Firstly, we sought to examine the cognitive profile of BDD as a diagnostic group as compared to healthy individuals in the largest sample to date using a comprehensive cognitive battery assessing eight cognitive domains: Working memory, speed of processing, reasoning and problem-solving, visual learning, verbal learning, attention/vigilance, social cognition, and inhibition/flexibility (12, 13). Based on the existing literature (5-10), we predicted that the BDD group would demonstrate significantly poorer performance than healthy controls in domains of inhibition/flexibility, speed of processing, reasoning and problem-solving, working memory, visual learning and attention/vigilance. Secondly, we aimed to explore whether disparate cognitive subgroups may be present within BDD, by subjecting participants’ cognitive data to hierarchical cluster analysis. Consistent with research in other psychiatric conditions (12) and the extant literature on cognitive functioning in BDD, we hypothesised that at least two distinct cognitive subgroups, exhibiting profiles ranging from intact to impaired cognitive performance, would be identified within the BDD group.

## Method

### Participants

The sample comprised 65 individuals with BDD and 70 healthy controls. Participants were recruited through treating psychiatrists or psychologists with speciality clinical services treating BDD and public advertising. BDD diagnoses were confirmed using the BDD Diagnostic Module for DSM criteria (14) and major psychiatric diagnoses were screened using the Mini International Neuropsychiatric Interview (MINI; 15). Individuals with BDD were excluded if BDD was not determined to be the current primary psychiatric diagnosis. At the time of testing, 77.8% of the BDD sample had at least one currently comorbid psychiatric disorder according to MINI screening. Comorbidities included major depressive disorder (*n =* 23, 42.6%), generalised anxiety disorder (*n =* 24, 44.4%), panic disorder (*n =* 11, 20.4%), social anxiety disorder (*n =* 16, 29.6%), obsessive-compulsive disorder (*n =* 4, 7.4%), post-traumatic stress disorder (*n =* 2, 3.7%), alcohol use disorder (within past 12 months, *n =* 9, 16.7%), substance use disorder (within past 12 months, *n =* 4, 7.4%), any psychotic disorder (*n =* 1, 1.9%; no psychotic symptoms within previous month at time of testing), anorexia nervosa (*n =* 2, 3.7%), and bulimia nervosa (*n =* 4, 7.4%). Comorbidity data was missing for 11 (16.9%) BDD participants. At the time of testing, 29 (44.6%) of BDD participants were taking psychiatric medications (see Online Supplement for medication details). All participants were stabilised on medication at time of testing (i.e., had not started a new medication or discontinued a medication within previous four weeks).

Healthy control participants were excluded if they screened positive to any current psychiatric diagnoses on the MINI or reported a history of significant mental illness. All participants were required to be at least 18 years of age, fluent in English, have an estimated premorbid IQ above 70 as scored using the Wechsler Test of Adult Reading (16), and no uncorrected hearing or visual impairments, neurological disorders or neurodevelopmental disorders. All research procedures contributing to this work were approved by hospital and university human research ethics committees (Alfred Hospital, St Vincent’s Hospital, Australian Catholic University, and Swinburne University of Technology) and complied with by the Declaration of Helsinki of 1975, as revised in 2008. Written informed consent was obtained from all participants before beginning study procedures.

### Clinical measures

The severity of BDD symptoms was assessed using the Yale-Brown Obsessive-Compulsive Scale Modified for BDD (BDD-YBOCS; 17). Illness insight was assessed using the Brown Assessment of Beliefs Scale (BABS; 18). The total score of the 42-item Depression, Anxiety, and Stress Scale (DASS; 19) was used to evaluate the severity of depression, anxiety, and stress symptoms over the previous week.

### Cognitive measures

The Measurement and Treatment Research to Improve Cognition in Schizophrenia (MATRICS) Consensus Cognitive Battery (MCCB; 13) was used to assess seven domains of cognitive functioning, as derived from scores on ten individual subtests: *i*) *Speed of processing*, derived from the Brief Assessment of Cognition in Schizophrenia Symbol Coding Test, the Category Fluency Animal Naming Test, and the Trail Making Test: Part A; *ii*) *Working memory*, derived from the Letter Number Span and Wechsler Memory Scale: Spatial Span; *iii*) *Reasoning and problem-solving*, derived from the Neuropsychological Assessment Battery Mazes; *iv*) *Visual learning*, derived from the Brief Visuospatial Memory Test – Revised; *v*) *Verbal learning*, derived from Hopkins Verbal Learning Test – Revised; *vi) Attention/vigilance*, derived from the Continuous Performance Test – Identical Pairs; and *vii) Social cognition*, derived from the Myer-Salovey-Caruso Emotional Intelligence Test: Managing Emotions branch. Two additional tests were used to form an eighth cognitive domain representing *Inhibition/flexibility* (12). Scaled scores from the inhibition condition of the Delis-Kaplan Executive Function System version of the Stroop Colour-Word Interference Test (20) were used as a measure of inhibition. The Trail Making Test: Part B (21) raw completion time was used as a measure of flexibility. All cognitive measures were administered in person at our research facility.

### Statistical analysis

In initial analyses, clinical and demographic characteristics were compared between BDD and control participants using one-way analyses of variance (ANOVA), or chi-square (χ^2^) tests of independence. Before proceeding with our primary analyses, missing cognitive data were replaced using EM imputation as performed in IBM SPSS v26. Next, cognitive data were reversed as necessary to ensure that higher scores always indicated better test performance. Cognitive data were then standardised to *z*-scores (*M =* 0, *SD* = 1) relative to the healthy control group’s means, and the eight cognitive domains were created by combining the relevant standardised subtest scores. To account for low performance that may nevertheless be within a normative range, we applied a liberal threshold of one standard deviation or more below the healthy control group’s average as a criterion for clinically low performance (22).

To achieve our first aim, performance of the BDD and healthy control groups were compared across each of the eight cognitive domains using ANOVAs (Bonferroni corrected α = .05/8 = .006). As age-related differences have been reported in the cognitive domains under assessment (23), age was included as a second-step covariate in all omnibus tests. To evaluate associations between state symptoms and cognitive performance, Pearson bivariate correlations were conducted among each cognitive domain score and DASS total score, BDD symptom severity (BDD-YBOCS), and illness insight (BABS) scores.

To achieve our second aim, hierarchical cluster analysis was performed on the BDD group’s cognitive data using Euclidean distances and Ward’s linkage method. Dendrogram inspection and examination of the agglomeration schedule and scree plot of agglomeration coefficients were used to inform the interpretation of cluster solutions (24). Cognitive domain scores of the resulting clusters were compared to each other and to healthy controls using ANOVAs with (Bonferroni corrected α = .05/8 =.006), followed up with Games-Howell post-hoc tests (α = .05/18 = .003). Cross-cluster comparisons of clinical and sociodemographic variables were performed using one-way ANOVA and χ^2^ tests (Bonferroni corrected α = .05/23 = .002). Finally, discriminant functions analysis was conducted to evaluate the validity of the clusters and investigate the relative contribution of cognitive domains to the final clustering solutions. Measures of effect size (Cohen’s *d*, omega-squared (ω^2^), or Cramer’s *V*) were calculated for all comparisons.

## Results

### Sample characteristics

The BDD and healthy control groups were statistically similar in mean age (*F*(1,133) = 1.90, *p =* .166, *d* = 0.24), estimated IQ (*F*(1,133) = 4.29, *p* = .040, *d* = 0.36), sex distribution (χ^2^(1) = 0.23, *p* = .635, *V* = 0.64) and current occupational status (χ^2^(3) = 6.50, *p* = .090, *V* = 0.22; see Table 1). BDD participants had significantly higher DASS total scores than healthy controls, *F*(1,133) = 96.67, *p* < .001, *d* = 1.90. Patterns of highest education differed significantly between the groups, with results indicating lesser educational attainment in BDD, χ^2^(4) = 20.08, *p* < .001, *V* = 0.39.

**Table 1.** Demographic and clinical characteristics in BDD and healthy control groups.

| *Characteristics* | Healthy controls<br>(*n* = 70) |  | BDD<br>(*n* = 65) |  |
| --- | --- | --- | --- | --- |
|  | *M* | *SD* | *M* | *SD* |
| Age in years | 30.2 | 9.1 | 32.9 | 12.7 |
| Estimated IQ | 110.8 | 7.5 | 106.9 | 13.5 |
| DASS Total <sup>†</sup> | 11.9 | 10.3 | 52.2 | 28.2 |
| BDD Severity | - | - | 26.8 | 7.6 |
| Illness insight | - | - | 14.0 | 6.3 |
| Age at onset <sup>‡</sup> | - | - | 19.6 | 10.1 |
| Illness duration <sup>‡</sup> | - | - | 13.6 | 10.6 |
|  | *n* | % | *n* | % |
| Females | 37 | 52.9% | 37 | 56.9% |
| *Highest educational attainment* |  |  |  |  |
| Primary school | - | - | 1 | 1.5% |
| Secondary school | 9 | 12.9% | 27 | 41.5% |
| Trade qualification, diploma or certificate | 12 | 17.1% | 14 | 21.5% |
| Undergraduate degree | 33 | 47.2% | 13 | 20.0% |
| Postgraduate degree | 16 | 22.9% | 10 | 14.4% |
| *Occupational status* <sup>§</sup> |  |  |  |  |
| Full time employment or study | 32 | 45.7% | 17 | 30.9% |
| Part-time employment and/or part-time study | 19 | 21.7% | 16 | 29.1% |
| Home or carer duties, or volunteering | 6 | 8.6% | 3 | 5.5% |
| Unemployed or retired | 13 | 18.6% | 19 | 34.5% |

*Note.* <sup>†</sup> Missing data for 29 cases; <sup>‡</sup> Missing data for four cases; <sup>§</sup> Missing data for 10 cases; *d* = Cohen's *d* effect size, small = 0.2, medium = 0.5, large = 0.8; *V* = Cramer's *V* effect size, 0 = no association, 1 = strong association.

### Cognition in BDD versus Controls

Main comparisons demonstrated significantly poorer performance in BDD relative to healthy controls across domains of inhibition/flexibility, speed of processing, working memory, verbal learning, visual learning, and reasoning and problem-solving (Table 2). The groups did not significantly differ in attention/vigilance or social cognition. When controlling for age, main effects for inhibition/flexibility were no longer significant according to the Bonferroni corrected threshold, *F*(1,132) = 7.33, *p* = .008). All other results remained unchanged. Cognitive domain scores were not significantly correlated with DASS total score in either group, or with BDD symptom severity and insight scores in the BDD group.

**Table 2.**
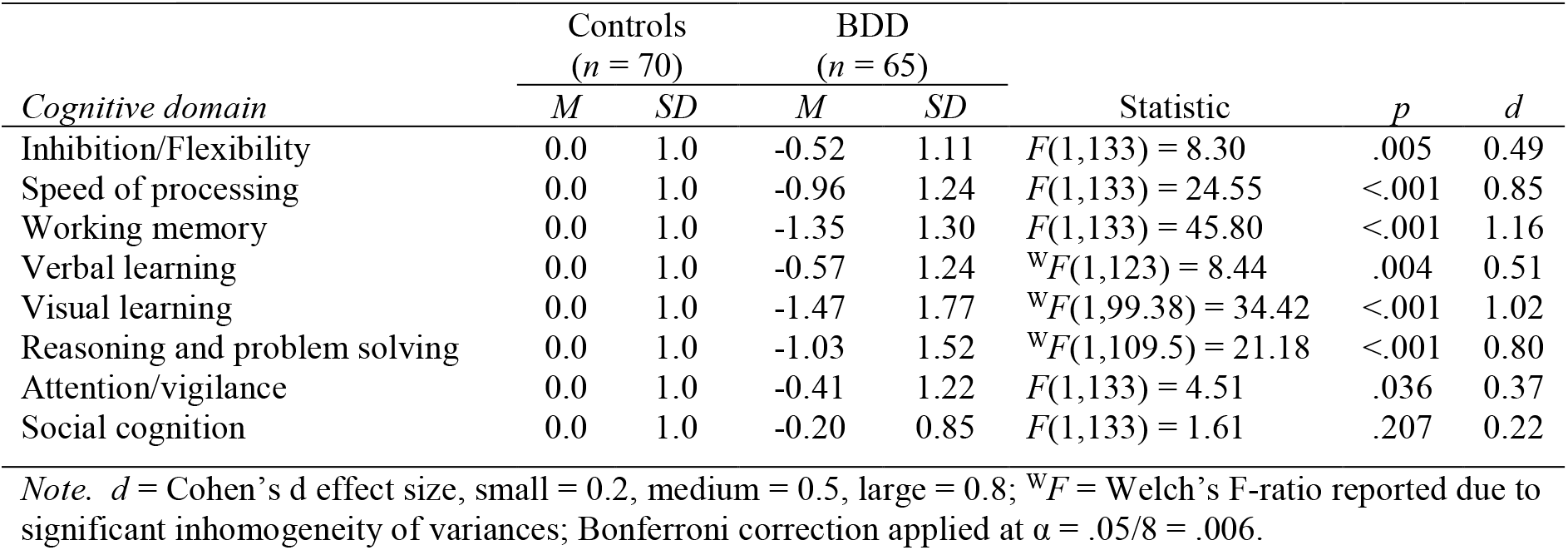
Group-wise cognitive domain and subtest performance as standardised to relative to the healthy control group.

| *Cognitive domain* | Controls<br>(*n* = 70) |  | BDD<br>(*n* = 65) |  | Statistic | *p* | *d* |
| --- | --- | --- | --- | --- | --- | --- | --- |
|  | *M* | *SD* | *M* | *SD* |  |  |  |
| Inhibition/Flexibility | 0.0 | 1.0 | -0.52 | 1.11 | $F(1,133) = 8.30$ | .005 | 0.49 |
| Speed of processing | 0.0 | 1.0 | -0.96 | 1.24 | $F(1,133) = 24.55$ | <.001 | 0.85 |
| Working memory | 0.0 | 1.0 | -1.35 | 1.30 | $F(1,133) = 45.80$ | <.001 | 1.16 |
| Verbal learning | 0.0 | 1.0 | -0.57 | 1.24 | $^wF(1,123) = 8.44$ | .004 | 0.51 |
| Visual learning | 0.0 | 1.0 | -1.47 | 1.77 | $^wF(1,99.38) = 34.42$ | <.001 | 1.02 |
| Reasoning and problem solving | 0.0 | 1.0 | -1.03 | 1.52 | $^wF(1,109.5) = 21.18$ | <.001 | 0.80 |
| Attention/vigilance | 0.0 | 1.0 | -0.41 | 1.22 | $F(1,133) = 4.51$ | .036 | 0.37 |
| Social cognition | 0.0 | 1.0 | -0.20 | 0.85 | $F(1,133) = 1.61$ | .207 | 0.22 |

*Note.* *d* = Cohen's *d* effect size, small = 0.2, medium = 0.5, large = 0.8; $^wF$ = Welch's *F*-ratio reported due to significant inhomogeneity of variances; Bonferroni correction applied at $\alpha = .05/8 = .006$ .

### Clustering analysis

A two-cluster solution was determined to provide the most appropriate fit for the BDD data. Figure 1 illustrates the agglomeration of cases leading to the final two-cluster solution and the cognitive profiles of each cluster. Significant differences were found among the two BDD clusters and healthy controls in all cognitive domains except attention/vigilance and social cognition (Tables 3 and 4). The pattern of significant results remained unchanged when controlling for age.

**Table 3.** Means and group comparisons of cognitive domain performance among the two BDD clusters and healthy controls.

| *Cognitive domain* | Controls<br>(*n* = 70) | | Cluster 1<br>(*n* = 18) | | Cluster 2<br>(*n* = 47) | | Statistic | *p* | $\omega^2$ |
| --- | --- | --- | --- | --- | --- | --- | --- | --- | --- |
|  | *M* | *SD* | *M* | *SD* | *M* | *SD* |  |  |  |
| Inhibition/flexibility | 0.0 | 1.0 | -1.51 | 1.12 | -0.15 | 0.86 | $F(2,132) = 17.80$ | <.001 | 0.47 |
| Speed of processing | 0.0 | 1.0 | -2.15 | 0.88 | -0.50 | 1.04 | $F(2,132) = 33.37$ | <.001 | 0.32 |
| Working memory | 0.0 | 1.0 | -2.55 | 1.05 | -0.89 | 1.08 | $F(2,132) = 45.33$ | <.001 | 0.40 |
| Verbal learning | 0.0 | 1.0 | -1.15 | 1.13 | -0.34 | 1.22 | $F(2,132) = 7.97$ | .001 | 0.09 |
| Visual learning | 0.0 | 1.0 | -3.40 | 1.67 | -0.73 | 1.15 | $^wF(2,41.4) = 35.85$ | <.001 | 0.47 |
| Reasoning and problem solving | 0.0 | 1.0 | -2.65 | 1.31 | -0.40 | 1.07 | $F(2,132) = 44.46$ | <.001 | 0.39 |
| Attention/vigilance | 0.0 | 1.0 | -0.79 | 1.02 | -0.26 | 1.27 | $F(2,132) = 3.78$ | .025 | 0.04 |
| Social cognition | 0.0 | 1.0 | -0.16 | 1.20 | -0.22 | 0.70 | $^wF(2,43.79) = 0.99$ | .381 | 0.00 |

*Note.* $\omega^2$ = Omega squared effect size, small = .01, medium = .06, large = .14; $^wF$ = Welch's *F* reported due to significant inhomogeneity of variances; Bonferroni correction applied at $\alpha = .05/8 = .006$ .

**Table 4.** Games-Howell post-hoc comparisons of cognitive domain scores across healthy controls and the two BDD clusters.

| *Cognitive domain* | *Post-hoc result* | *p* | *d* |
| --- | --- | --- | --- |
| Inhibition/flexibility | Controls > Cluster 1 | <.001* | 1.42 |
|  | Controls = Cluster 2 | .676 | 0.16 |
|  | Cluster 1 < Cluster 2 | <.001* | 1.00 |
| Speed of processing | Controls > Cluster 1 | <.001* | 2.28 |
|  | Controls = Cluster 2 | .030 | 0.49 |
|  | Cluster 1 < Cluster 2 | <.001* | 1.71 |
| Working memory | Controls > Cluster 1 | <.001* | 2.71 |
|  | Controls > Cluster 2 | <.001* | 0.86 |
|  | Cluster 1 < Cluster 2 | <.001* | 1.56 |
| Verbal learning | Controls > Cluster 1 | .002* | 1.08 |
|  | Controls = Cluster 2 | .250 | 0.30 |
|  | Cluster 1 = Cluster 2 | .044 | 0.69 |
| Visual learning | Controls > Cluster 1 | <.001* | 2.47 |
|  | Controls > Cluster 2 | .002* | 0.68 |
|  | Cluster 1 < Cluster 2 | <.001* | 1.86 |
| Reasoning and problem solving | Controls > Cluster 1 | <.001* | 2.27 |
|  | Controls = Cluster 2 | .106 | 0.39 |
|  | Cluster 1 < Cluster 2 | <.001* | 1.88 |

*Note.* \*Significant at Bonferroni corrected $\alpha = .05/18 = .003$ ; $d$ = Cohen's $d$ effect size. Relevant omnibus test results are reported in Table 3. Post-hoc tests for attention/vigilance and social cognition domains were not performed due to insignificant omnibus tests.

**Figure 1.**
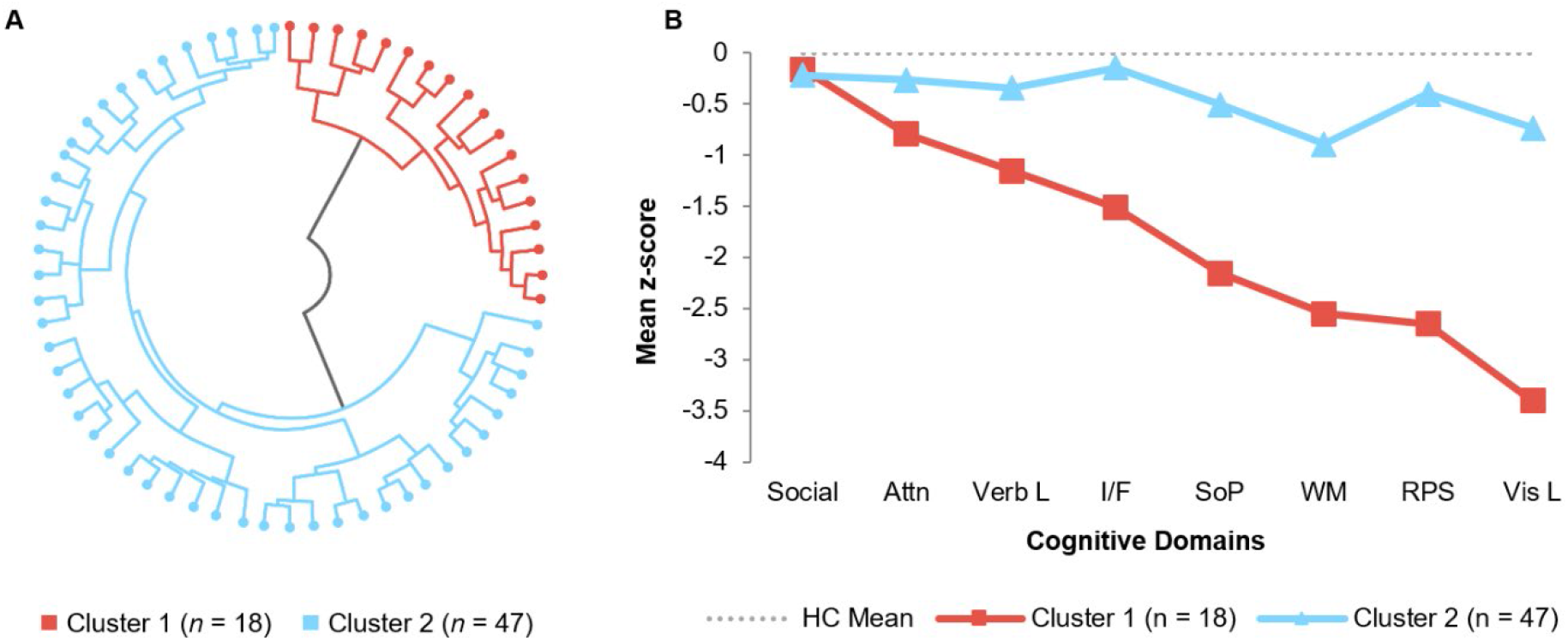
Results of hierarchical clustering analysis in BDD. (A) The radial dendrogram illustrates the obtained pattern of hierarchical agglomerative clustering, resulting in the determination of two BDD clusters. Each dot on the outer circumference represents a single BDD case, with connecting lines representing the combining of cases to form clusters. (B) The line graph shows cognitive profiles of the two BDD clusters plotted according to mean cognitive domain z-scores, as standardised to the healthy control group. HC = Healthy control, Social = Social cognition, Attn = attention/vigilance, Verb L = verbal learning, I/F = inhibition/flexibility, SoP = speed of processing, WM = working memory, RPS = reasoning and problem-solving, Vis L = visual learning. *[Note: Greyscale figure also available for printed issues]*.

Cluster 1 (*n* = 18) was characterised by significantly poorer scores than healthy controls for all cognitive domains except for attention/vigilance and social cognition. Cluster 1 further demonstrated significantly poorer inhibition/flexibility, speed of processing, working memory, visual learning, and reasoning and problem-solving in comparison to Cluster 2. Cluster 2 (*n* = 47) was characterised by significantly poorer working memory and visual learning as compared to healthy controls. There were no further significant differences between Cluster 2 and healthy controls in the remaining domains. Based on these results, Cluster 1 was labelled as *broadly impaired*, and Cluster 2 was labelled as *broadly intact with mild selective impairments*.

Comparison of clinical and sociodemographic characteristics revealed a significantly higher mean age in Cluster 1 (*M* = 43.11, *SD* = 15.62) than in Cluster 2 (*M* = 28.89, *SD* = 8.68, *Welch’s F*(1, 21.2) = 13.34, *p* = .001, *d* = 1.13). No further significant differences were found for BDD symptom severity scores, insight scores, age at BDD onset, DASS total scores, estimated IQ, medication status, current comorbid diagnoses, sex distribution, current employment status or highest level of completed education (see Supplementary data).

### Discriminant functions analysis

One function accounting for 100% of variance significantly discriminated the clusters, Wilk’s λ = .270, χ^2^ (8) = 77.17, *p* < .001, Canonical correlation = .85. Structure matrix correlations indicated that the function was most strongly loaded on to by visual learning (*r =* .57), reasoning and problem-solving (*r* = .55), speed of processing (*r* = .46), working memory (*r* = .43) and inhibition/flexibility (*r* = .40). Negligible correlations were found for verbal learning (*r* = .19), attention/vigilance (*r* = .12), and social cognition (*r* = −.02). The function correctly classified cluster membership for 96.9% of BDD cases.

## Discussion

This study represents the largest and most comprehensive investigation of cognition in BDD to date. We examined the cognitive profile of BDD relative to healthy controls across eight domains and explored whether divergent cognitive clusters may be present within the BDD group. At the level of group-average comparisons, we found the BDD group had significantly lower scores than healthy controls in domains of inhibition/flexibility, speed of processing, working memory, verbal learning, visual learning, and reasoning and problem-solving with large effect sizes, especially for working memory and visual learning. However, no significant group differences were found for attention/vigilance or social cognition, or for inhibition/flexibility after controlling for age. Use of cluster analysis further revealed two distinct clusters within the BDD sample, characterised by cognitive performances ranging from *broadly intact with mild selective impairments* (referred to as *selectively impaired* hereafter for brevity; 72.3%) to *broadly impaired* (27.7%) cognitive functioning. The *selectively impaired* cluster demonstrated significantly poorer working memory (*M* = −0.89) and visual learning (*M* = −0.73) than healthy controls, despite showing mean scores within one standard deviation of the healthy control’s means on all cognitive domains. The *broadly impaired* cluster demonstrated significantly poorer performance than controls in all domains except social cognition and attention/vigilance, with very large effect sizes. As such, it is likely that the overall pattern of group-level differences between BDD and healthy control participants across many of the domains were largely driven by this subgroup.

These findings challenge contemporary views of cognition in BDD. Firstly, the results suggest that while people with BDD demonstrate cognitive difficulties at a diagnostic group-level, it cannot be assumed that the disorder is uniformly associated with cognitive deficits. Instead, our results demonstrate that there is considerable cognitive variation among persons with BDD. Yet the presence of these differing cognitive subgroups as identified via cluster analysis was masked in our group-level comparison of BDD to controls, with the severe magnitude of impairment in the *broadly impaired* subgroup being mitigated, and the presence of the *selectively impaired* subgroup altogether obviated. Thus, taking a diagnostic group-level approach may be detrimental to both research advances and understanding of cognition for individuals presenting with BDD. Secondly, the current results may help explain previous inconsistent findings, which have demonstrated both impaired and intact cognition in BDD relative to healthy controls across domains of inhibition, planning, visual and verbal learning, attention, working memory, and speed of processing (e.g., 5, 6, 7-11). Speculatively, we suggest that these mixed findings might stem from variations in the presence of differing cognitive subgroups across study samples.

However, visual learning and working memory were highlighted as areas of reduced performance across both BDD subgroups, despite the *broadly impaired* subgroup demonstrating significantly poorer scores in these domains as compared to the *selectively impaired* subgroup. These findings may suggest a generalised vulnerability to visual learning and working memory difficulties in BDD, though the degree of deficit may vary widely across individuals. Indeed, visual learning emerged as the strongest contributing factor to the discrimination of clusters, closely followed by reasoning and problem-solving, speed of processing, working memory and inhibition/flexibility. These results indicate that differences in visual and ‘executive’ functions (i.e., encompassing working memory, attention, reasoning and problem-solving, inhibition/flexibility) may be important in demarcating divergent cognitive subgroups within the disorder. However, the inter-relationships between these functions requires further investigation. It has previously been suggested that executive dysfunction may underlie poor performance in other cognitive domains in BDD, including visual learning (9). An additional interpretation might be that disrupted visual processing could impair performance on visually-based executive tasks (e.g., mazes, spatial span). Specifically, atypical dominance of detail-oriented local visual processing over global processing has been repeatedly found in studies of BDD, while neuroimaging data has shown aberrant visual stream activation in the disorder (25, 26). To tease apart potential interactions of visual and executive functions in BDD, further research which contrasts visual and non-visual measures of executive functioning is necessary.

Our data suggest that age may have had some influence on the obtained results, though any such influence is likely to be circumscribed. The *broadly impaired* subgroup demonstrated a significantly older mean age than the *selectively impaired* subgroup, and inhibition/flexibility was not significantly different between the overall BDD sample and healthy controls after controlling for age (despite a medium effect size for group differences). In healthy samples, participants aged younger than 50 years tend to show better performance on the MCCB than do older participants, except in domains of social cognition and verbal learning (23). Though the mean age of the *broadly impaired* cluster was approximately 43 years, the dispersion of age was broad (ranging from 19 to 62 years). The age range for the *selectively impaired* cluster was also broad (18 to 49 years), and co-varying for age did not lead to any negation of the significance of between-cluster differences across cognitive domain performances. As such, the poor performance of the *broadly impaired* subgroup is unlikely to be attributable to age alone.

Importantly, we found no significant differences in all remaining sociodemographic or clinical features between the two BDD clusters. Moreover, cognitive domain performances did not significantly correlate with BDD symptom severity, insight, or DASS total score in the broader BDD group. These findings imply a dissociation between illness characteristics and cognitive functioning, which has implications for psychological models of BDD that propose an underlying role of cognitive deficits in the development or maintenance of symptoms (4). Further research is warranted to replicate these findings, and to understand the potential basis of cognitive changes in BDD, including why some individuals demonstrate marked and widespread impairments while others do not. This could include mapping cognitive profiles on to neurobiological substrates (26), or longitudinal approaches which test the stability of cognitive subgroups across the lifespan or illness course.

Social cognition was intact in both BDD clusters. This finding is at odds with prior reports of inaccurate social inferences and poor recognition of facial expressions in BDD (27, 28). This discrepancy may stem from differences in the precise social cognitive constructs being assessed across studies. In particular, the MCCB subtest for social cognition, as used presently, assesses the application of an intellectual understanding of emotions to decision-making in hypothetical social scenarios (13). Conversely, noted impairments in BDD have involved other aspects of social cognition, such as recognition of emotional facial expressions (28, 29), mentalising and decoding unspoken social cues (27), and biased social interpretations and attributions (30). It has been proposed that social cognitive impairments in BDD are circumscribed to aspects that may be influenced by negative, socially-oriented beliefs (e.g., risk of social threat), rather than intellectual aspects of understanding social processes (27). Accordingly, our findings suggest that intellectual aspects of social cognition are intact in BDD. Clearly, further research which contrasts these different aspects of social cognition in BDD is needed.

Finally, our dual-cluster solution in BDD represents a different structure of within-disorder cognitive heterogeneity to that which has been established in schizophrenia and bipolar disorder, which has typically indicated at least a three-cluster pattern (i.e., intact, mild-moderately impaired, and globally impaired subgroups; 12). Moreover, the proportion of individuals with broadly impaired cognition appears smaller in BDD than in these disorders (12), and the specific cognitive profiles of our obtained BDD clusters also appear qualitatively different. Specifically, our results demonstrate greater visual learning impairment in our *broadly impaired* BDD cluster, as compared to equivalent schizophrenia/bipolar subgroups (wherein speed of processing, attention/vigilance, and working memory were most impaired; 12). As such, our data might suggest that there are underlying differences in cognitive phenotypes between BDD and these disorders, despite sharing other similarities such as equivalent endorsement of delusional themes related to control and thought alienation (31). Comparison of cognitive subgroups in BDD as relative to other disorders may thus be informative in determining the specificity of observed cognitive phenotypes, particularly in conceptually related disorders such as obsessive-compulsive disorder (32).

The current study represents the largest investigation of cognition in BDD to date and is the first to examine cognitive heterogeneity within BDD. The use of an established battery, which spans multiple domains of cognitive functioning, further represents a significant strength, as does the use of a well-matched healthy control group from which an appropriate point of reference was derived. However, there are some caveats. The output of clustering analysis is dependent on choice of variables entered, and therefore the choice of tests selected for measurement of cognitive function. Selection of different tests might have produced a different pattern of cluster solutions or profiles. Future research may investigate the reproducibility of obtained cluster patterns by replicating the current analysis with analogue tests. In addition, although we recorded and examined the role of medication status and comorbidities in this study, these factors could exert a confounding influence that future studies might aim to address.

In sum, this work provides novel evidence of cognitive heterogeneity in BDD, alongside the largest group-level comparison of cognition in BDD relative to healthy controls to date. The finding of two divergent cognitive subgroups in BDD characterised by *selectively impaired* and *broadly impaired* cognitive functioning highlights the need to consider a nuanced approach in studies of cognition in BDD. However, as we found little significant differences in clinical or sociodemographic variables among these cognitive clusters, the possible role of cognitive functioning in the aetiology or clinical presentation of BDD remains unclear. Future research should aim to investigate mechanisms which might explain such divergences of cognitive functioning among individuals with BDD, perhaps with neurobiological or longitudinal approaches.

## Data Availability

The data is not available for sharing.

## Acknowledgements

A.M, S.B, S.A.G, and T.P were supported by Australian Government Research Training Program Scholarships during this research. W.L.T holds a National Health and Medical Research Council (NHMRC) grant (GNT1161609). S.L.R holds a Senior NHMRC Fellowship (GNT1154651). This study was funded in part by a Monash Strategic Grant, a Barbara Dicker Brain Sciences Foundation Grant, and an Australian Catholic University Research Fund grant (ACURF2013000557).

